# Estimating the establishment of local transmission and the cryptic phase of the COVID-19 pandemic in the USA

**DOI:** 10.1101/2020.07.06.20140285

**Authors:** Jessica T. Davis, Matteo Chinazzi, Nicola Perra, Kunpeng Mu, Ana Pastore y Piontti, Marco Ajelli, Natalie E. Dean, Corrado Gioannini, Maria Litvinova, Stefano Merler, Luca Rossi, Kaiyuan Sun, Xinyue Xiong, M. Elizabeth Halloran, Ira M. Longini, Cécile Viboud, Alessandro Vespignani

## Abstract

We use a global metapopulation transmission model to study the establishment of sustained and undetected community transmission of the COVID-19 pandemic in the United States. The model is calibrated on international case importations from mainland China and takes into account travel restrictions to and from international destinations. We estimate widespread community transmission of SARS-CoV-2 in February, 2020. Modeling results indicate international travel as the key driver of the introduction of SARS-CoV-2 in the West and East Coast metropolitan areas that could have been seeded as early as late-December, 2019. For most of the continental states the largest contribution of imported infections arrived through domestic travel flows.

---

The first confirmed case of COVID-19 in the United States (US) was diagnosed in Washington state on January 21, 2020 (*1*). In quick succession other cases were confirmed in Illinois, California, and Arizona (*2-4*). All initial cases reported in the US had a known travel history to mainland China, the epicenter of the pandemic. The first reported *local* transmission on US soil was discovered on January 30 in Illinois (*5*). However, very few COVID-19 cases were detected until the case definition for testing was updated on March 4, 2020, to include symptomatic individuals without travel history (*6*). On April 1, when the US federal government issued social distancing guidelines, 26,655 new reported cases and 1,050 deaths were recorded in the United States. In Fig. 1A we show a timeline of initial confirmed cases, testing policies, and early containment and mitigation initiatives concerning the US COVID-19 outbreak.

**Figure 1:**
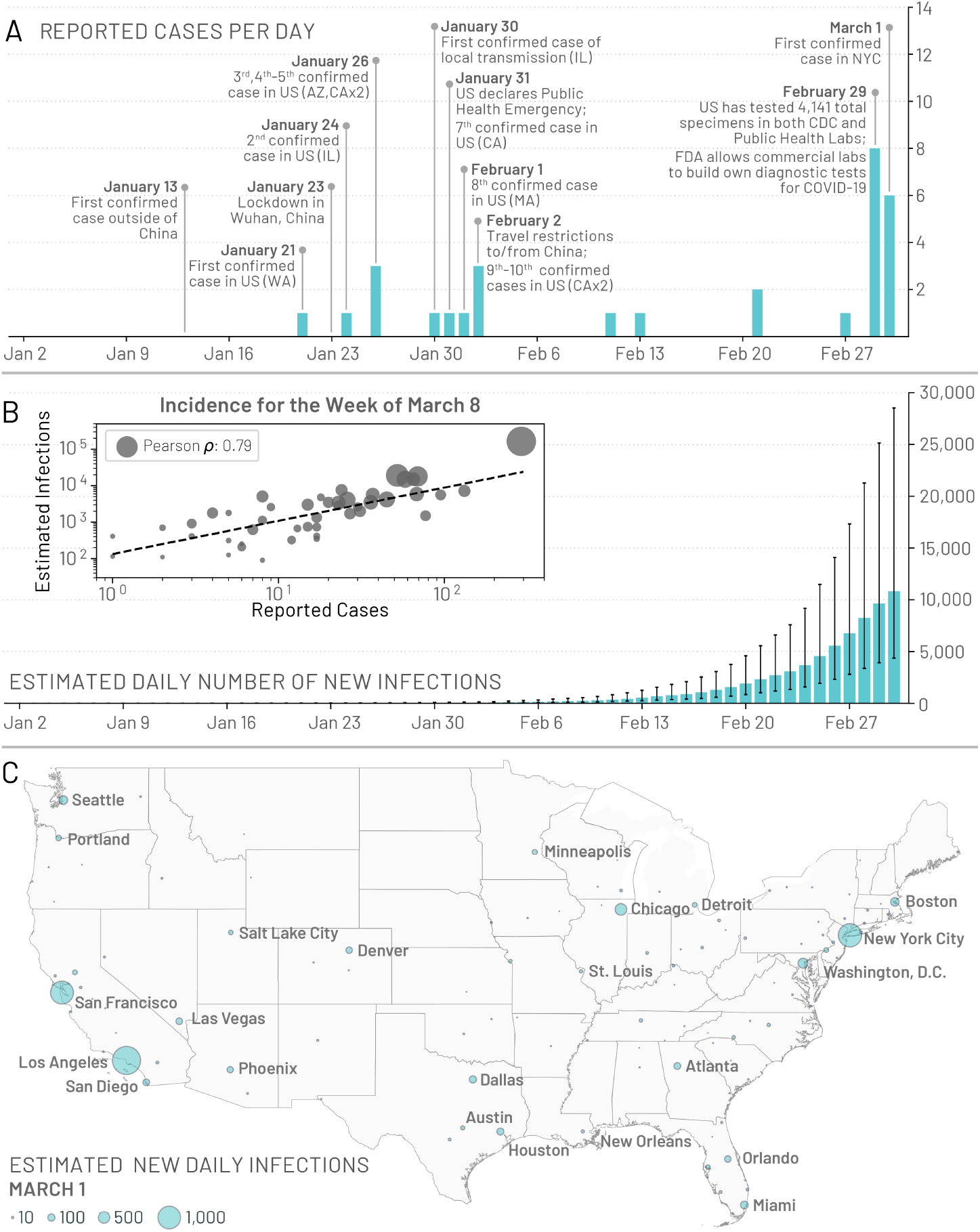
Early picture of the COVID-19 outbreak in the United States. (A) A timeline of the daily reported and confirmed cases of COVID-19 in the US including information on the first 10 reported cases and other significant events related to the outbreak up to March 1, 2020. (B) Model-based estimates for the median daily number of new infections in the continental US. Error bars represent the interquartile range. The inset plot compares the weekly incidence of reported cases with the weekly incidence of infections estimated by the model for the week of March 8 – 14, 2020, for 48 of the continental states that reported at least 1 confirmed case. Circle size corresponds to the population size of each state. (C) Model based estimates for the median number of daily infections in the continental US as of March 1, 2020.

Given the narrowness of the initial testing criteria, it is expected that the SARS-CoV-2 virus causing COVID-19 was spreading through “cryptic transmission” in January and February, setting the stage for the large epidemic wave experienced in March and April, 2020. In this study, we model the arrival and cryptic phase of the COVID-19 epidemic in the US. We estimate the time frame for the establishment of local transmission in the different states and provide a statistical analysis of the domestic spread of the COVID-19 epidemic.

To study the spatial and temporal spread of COVID-19, we use the Global Epidemic and Mobility Model (GLEAM), an individual-based, stochastic, and spatial epidemic model (*7-11*). The model was previously used to characterize the early stage of the COVID-19 epidemic in mainland China and the effect of travel restrictions on infections exported to other global regions (*12*). GLEAM generates an ensemble of possible epidemics described by the number of newly generated infections, the time of disease arrival in different regions of the world, and the number of infected travelers. The model divides the global population into more than 3, 200 subpopulations in roughly 200 different countries and territories. The airline transportation data encompass daily origin-destination traffic flows from the Official Aviation Guide (OAG) and the International Air Transport Association (IATA) databases (*13, 14*), whereas ground mobility flows are derived from the analysis and modeling of data collected from the statistics offices of 30 countries on five continents (*7, 8*).

The transmission dynamics take place within each subpopulation and assume a classic SLIR-like compartmentalization scheme for disease progression similar to those used in several large scale models of SARS-CoV-2 transmission (*15–20*). Each individual, at any given point in time, is assigned to a compartment corresponding to their particular disease-related state (being, e.g., susceptible, latent, infectious, removed) (*12*). This state also controls the individual’s ability to travel (details in the supplementary material, SM). Individuals transition between compartments through stochastic chain binomial processes. Susceptible individuals can acquire the virus through contacts with individuals in the infectious category and can subsequently become latent (i.e., infected but not yet able to transmit the infection). The process of infection is modeled using age-stratified contact patterns at the state level (*21*). Latent individuals progress to the infectious stage at a rate inversely proportional to the latent period, and infectious individuals progress to the removed stage at a rate inversely proportional to the infectious period. The sum of the mean latent and infectious periods defines the generation time. Removed individuals are those who can no longer infect others. To estimate the number of deaths, we use the age-stratified infection fatality ratios from (*22*). At this stage, the transmission model does not account for heterogeneities due to age differences in susceptibility to the SARS-CoV-2 infection. This is an intense area of discussion at the moment (*25–27*).

We assume a start date of the epidemic in Wuhan, China, that falls between November 15, 2019 and December 1, 2019, with 20 initial infections (*12, 20, 28–31*) (see SM for sensitivity analysis). The model generates an ensemble of possible epidemic realizations and is calibrated using Approximate Bayesian computation (ABC) methods (*32*) based on the observed international importations from mainland China through January 21, 2020 (*12*). Only a fraction of imported cases are detected at the destination (*33*). According to the estimates proposed in (*34*), we stratify the detection capacity of countries into three groups: high, medium and low surveillance capacity according to the Global Health Security Index (*35*), and assume asymptomatic infections are never detected (see SM). The model calibration does not consider correlated importations (family travel) and assumes that travel probabilities are homogeneous across all individuals in each subpopulation.

The ABC calibration using a generation time *T_g_* = 6.5 days yields 3, 700 individual realizations of the global evolution of the pandemic that provide information on imported infections, locally generated infections, and deaths in all subpopulations considered in the model (*36*). The model accounts for international travel restrictions according to available data on traffic reduction and government issued policies (See SM). The ABC calibration also gives a posterior distribution for the basic reproductive number *R*_0_ in the US (median 2.7 [95% CI 2.4-3.1]). The median reproductive numbers for each state range from 2.6 – 2.8, with doubling times ranging from 3.3 – 3.6 days (values are calculated using the the specific age stratified contact patterns for each state). In the SM we also consider an additional calibration based on the deaths observed in the US at the end of March. These results do not exhibit major differences and do not change the overall picture presented here.

## Onset of local transmission

Stochastic simulations of the worldwide epidemic spread yield international/domestic infection importations, incidence of infections, and deaths per subpopulation at a daily resolution in the continental US. In Fig.1B we show the model estimates for the median daily incidence of new infections up to March 1, 2020 in the continental US. There is a stark contrast between the model output and the number of officially reported cases by the same date, highlighting the significant number of transmission events that may have already occurred before many states had implemented testing strategies independent of travel history. For model validation we compare our projections of the number of infections during the week of March 8, 2020, to the number of cases reported during that week within 48 of the continental states that reported at least 1 case (shown in Fig.1B inset). We see a strong correlation between the reported cases and our model’s projected number of infections, (Pearson’s correlation coefficient on log-values 0.79, *p* < 0.001), although many fewer cases had actually been reported by that time. If we assume that the number of reported cases and simulated infections are related through a simple binomial stochastic sampling process, we find that on average 5 in 1,000 infections (90%CI [1 – 25 per 1,000]) were detected by March 8, 2020. The ascertainment rate grows quickly as testing capacity increases, and our estimate doubled to detecting 11 in 1,000 infections (90%CI [2 – 36 per 1, 000]) by March 15, 2020. By April 1, 2020 our model suggests a detection rate of 96 cases per 1,000 infections (90%CI: [20- 289 per 1,000]). The Centers for Disease Control and Prevention (CDC) estimates that 42 to 166 in 1,000 infections were detected during March and April, 2020, at different locations in the US, with estimated numbers of infections at least 10 times greater than the number of reported cases in locations across seven different states (*37, 38*). These SARS-CoV-2 infections are also distributed heterogeneously across the US. In Fig.1C we show the model-based median daily number of new infections on March 1, 2020.

In Fig. 2 we plot the posterior distribution for the earliest date within each state when at least 10 new infections per day occurred in the community. California, New York and New Jersey are the first states with a probability larger than 50% to have experienced local transmission by mid January (California) or beginning of February (New York and New Jersey), 2020. However, the wide distribution of dates suggests that SARS-CoV-2 cryptic transmission may have started as early as December, 2019. The posterior distribution for the timing of the onset of local transmission peaks by mid February in Florida, Illinois, Maryland, Massachusetts, Texas, Washington, Arizona, Colorado, Connecticut, Georgia, Indiana, Michigan, Minnesota, Nevada, New Hampshire, North Carolina, Ohio, Oregon, Pennsylvania, Tennessee, and Utah Virginia. From the posterior distribution of Fig. 2, all states have a median date of the onset of local transmission by early March, with the large majority of them in February, 2020, a critical month for the cryptic spread of SARS-CoV-2 in the continental US. However, during that time, testing in the US was still focused on returning travelers from China.

**Figure 2:**
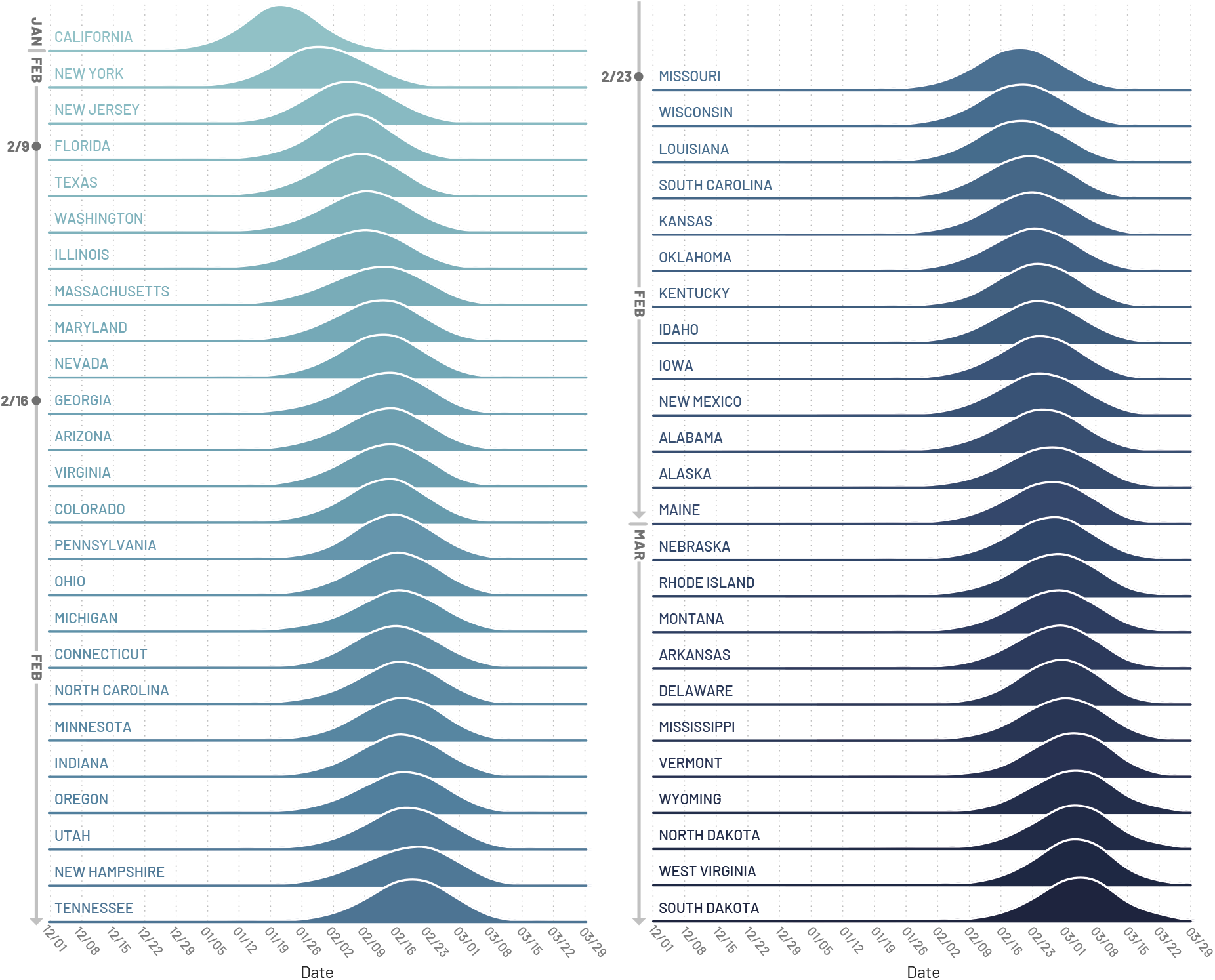
Timing of the onset of local transmission. Posterior distributions of the week when each state first reached 10 locally generated SARS-CoV-2 transmission events per day.

## International and domestic seeding

As the model allows the recording of the origin and destination of SARS-CoV-2 carriers at the global scale, we can study the possible sources of infection importation for each state. In particular, we are able to record the flow of latent and infectious individuals through international and domestic flight connections. However, the model also considers the effect of possible infections through commuters that may spend a few hours in a neighboring subpopulation (*7*). It is important to stress that the model’s realizations explore the many possible paths of the epidemic. Thus, the analysis provided here must be considered as a statistical description of the potential sources of SARS-CoV-2 importations, rather than providing a specific, single causal chain of events.

To identify seeding importation events relevant to the onset of local transmission in each stochastic realization of the model, we record the number of importations (of latent and infectious individuals) before the local transmission chains were established (defined as 10 daily local transmission events). We visualize the origin of the seeding importations relevant for establishing local transmission by aggregating the importation sources, considering some key geographical regions (e.g. Europe and Asia) while keeping the US and China separate and aggregating all the other countries (i.e. Others) in Fig. 3. It is worth clarifying that seeding importations are different from the actual number of times the virus has been introduced to each state with subsequent onward transmission. Even after local transmission has started, future importation events may give rise to additional onward transmission forming independently-introduced transmission lineages of the virus as seen in the United Kingdom (*39*). Statistics for importations through March 1, 2020, are reported in the SM file.

**Figure 3:**
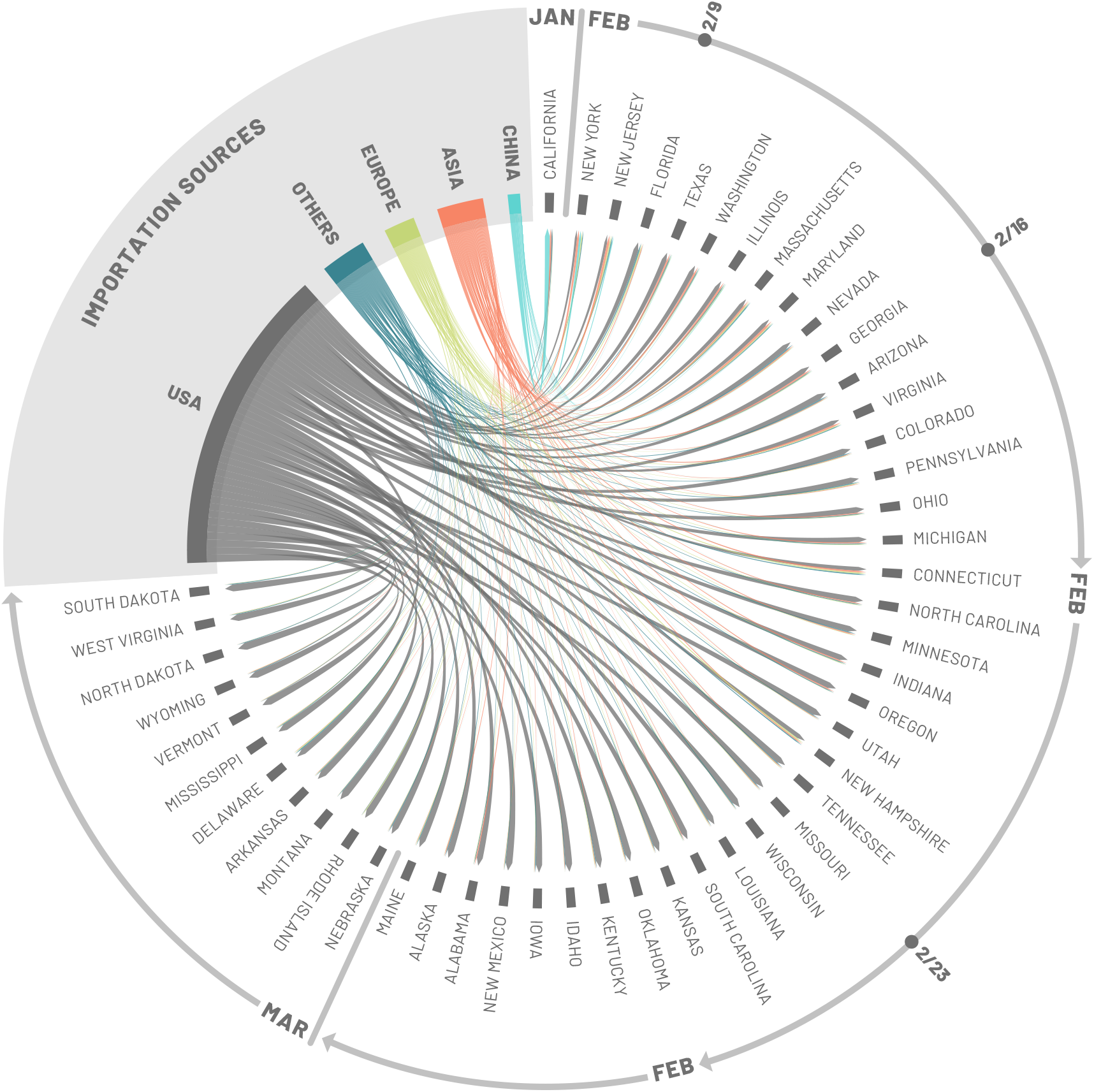
Importation sources. Each state is displayed in a clockwise order with respect to the start of the local outbreak (as seen in Fig. 2). Importation flows are directed and weighted. We normalize links considering the total in-flow for each state so that the sum of importations flows, for each state, is one. In the SM we report the complete list of countries contributing, as importation sources, in each group (i.e., geographical region).

Importations from mainland China may be relevant in seeding the epidemic in January (notice the width of the blue arrows from China for the first couple of states), but then play a small role in the COVID-19 expansion in the US because of the travel restrictions imposed to/from mainland China after January 23, 2020. About 60% and 24% of the virus introductions before the onset of local transmission in California and New York State, respectively, were from mainland China. While importations from mainland China contribute to early introductions of the virus in the US, our analysis highlights other potential sources of importations, such as Europe, where additional travel advisories and restrictions were implemented a month later; i.e. the end of February and early March. Noticeably, the share of infection importations originating from Europe in California is estimated to be about seven times smaller than those in New York State. Among the states for which the model estimates an early onset of local transmission before the second week of February (considering median values), European sources are statistically contributing 13% of SARS-Cov-2 importations for Florida, 16% for New Jersey, and only 5% for Washington. Interestingly, the domestic importations are, across the board, statistically relevant in seeding the epidemic in many states. Among the states for which we estimated a late onset of local transmission (second half of February or first week of March), domestic sources account for 77% of the virus introductions in Utah, 84% in New Mexico, 81% in Arkansas, and 90% in North Dakota.

## Cryptic spreading phase

From late January to early March, SARS-CoV-2 had been spreading across the US mostly undetected. We estimate that cities such as Los Angeles, New York, Chicago, Seattle, and San Francisco have possibly experienced local transmission beginning in the first half of February (Fig. 4A). In the time span of one additional month, through March 15, 2020, most large cities in the US had sizeable ongoing outbreaks. California most likely generated 10 local infections by late January (see Fig. 2), around the same time as the adoption of draconian containment measures and international travel restrictions in mainland China. The model also allows us to estimate possible COVID-19 related deaths. By March 1, 2020, we estimate a median of 63 [90% CI10 – 629] cumulative deaths in the US while only 1 death was reported. Although some states launched investigations in search of evidence that COVID-19 was the cause of death as far back as December 2019, it is likely that most deaths were missed because COVID-19 testing guidelines were based on travel history (*40*).

**Figure 4:**
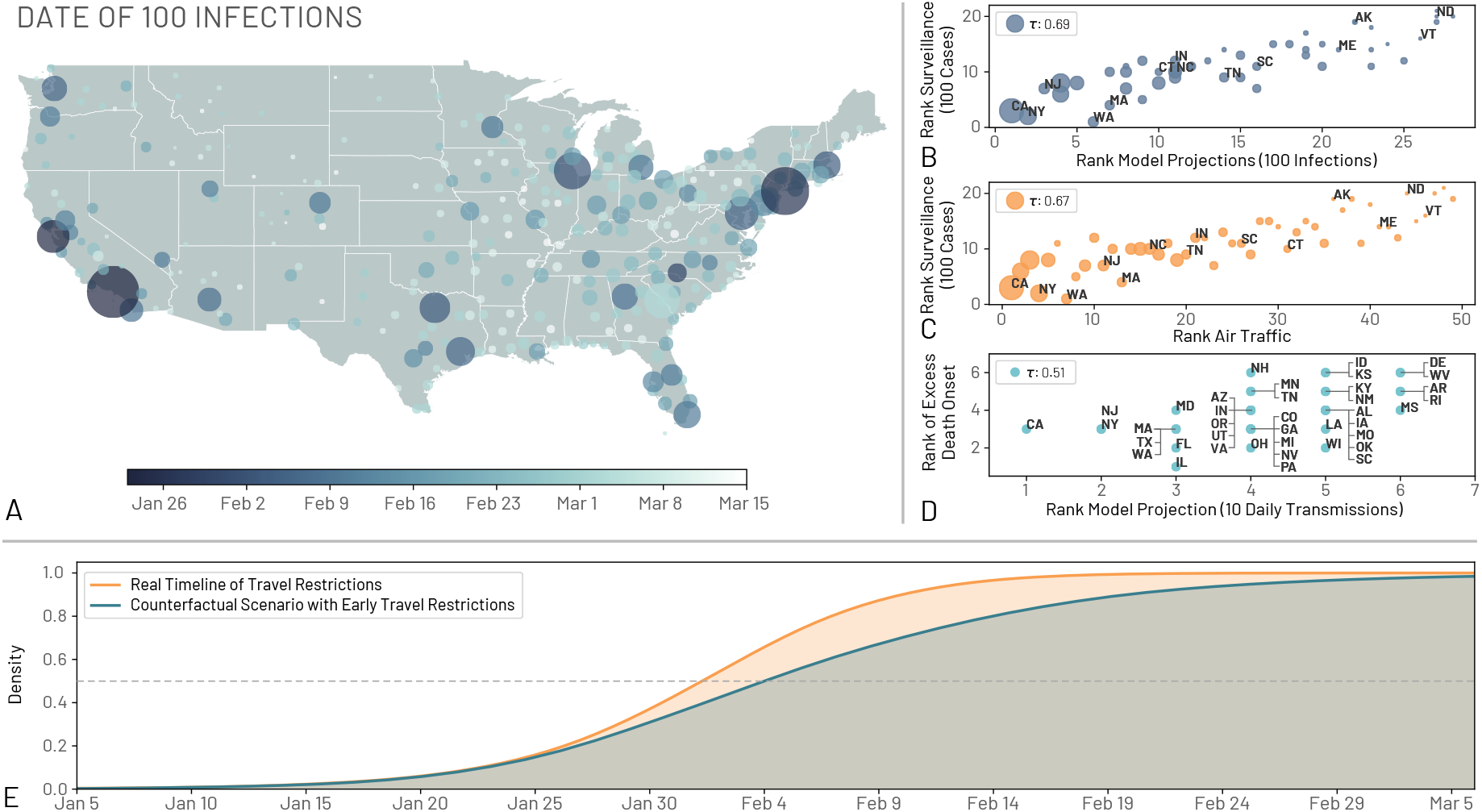
Comparing model projections to surveillance and air traffic data. (A) Map of the US showing the date where regions observed at least an average of 100 infections. (B) The correlation between the ordering of each state to reach 100 infections in the model projections and to reach 100 reported cases in the surveillance data. The correlation is computed using the Kendall rank correlation coefficient, *τ*. (C) The correlation between the ordering of each state considering the time needed to reach 100 reported cases in the surveillance data and the ranking of the combined international and domestic air traffic. (D) The order correlation of the onset of excess deaths due to pneumonia and influenza and the order of the model projection for the date of 10 transmissions per day. Circle size in A, B, and C corresponds to the population size of each state. (E) The cumulative distribution of the probability that the US reached 100 locally generated transmissions per day by a given day for both a scenario using the real timelines of travel restrictions and a counterfactual scenario where the travel restrictions to/from mainland China are shifted one week before. The median dates for the real and counterfactual timelines are February 2, 2020, and February 4, 2020, respectively. We show a horizontal line at a density of 0.5

To provide a model consistency check with respect to surveillance data concerning the epidemic, we compared the model-based estimates with observed surveillance data. Particularly, early on in the epidemic, surveillance data were known to be highly unreliable because of under-detection. For each state we compared the order in which they surpassed 100 infections in the model and in the surveillance data gathered from the John Hopkins University Coronavirus Resource Centre (*41*). In Fig. 4B we plot the ordering for states and compute the Kendall rank correlation coefficient *τ* (see SM for details). The correlation is positive (*τ* = 0.69, *p* < 0.001) indicating that, despite the detection and testing issues, the expected patterns of epidemic diffusion of the states is largely described by the model. As mentioned before, one possible major driver of the diffusion pattern is air traffic. We compare the ordering of states according to their air travel volume to their epidemic order as previously defined (Fig. 4C). We consider both national and international traffic and find a positive correlation (*τ* = 0.67 with *p* < 0.001) between the epidemic ordering derived from surveillance data and air traffic, suggesting the passenger volume of both international and national traffic are key factors driving the early phases of outbreak across the country. Similar observations have been reported in China, where the initial spreading of the virus outside Hubei was strongly correlated with the traffic to/from the province (*42*). The correlation between the air travel ordering and the simulations is high (*τ* = 0.87, *p* < 0.001) as actual airline data are used in the model to simulate the mobility of individuals (see SM). It is worth remarking that population size is also correlated with both the traveling flows (*τ* = 0.7, *p* < 0.001) and the epidemic order of each state (*τ* = 0.68, *p* < 0.001) as discussed in the SM. In our model it is not possible to exclude increased contacts in highly populated places before social distancing interventions and disentangle this effect from increased seeding due to the correlation between travel volume and population size. As yet another independent test for the model, in Fig. 4D we show the positive correlation between excess deaths, as estimated in (*43*), and the order in which the states reached the threshold of 10 daily transmissions (*τ* = 0.51, *p* < 0.001).

The model is consistent with the picture emerging from the genomic epidemiology (*44–46*) of an early start of the COVID-19 epidemic in coastal states, followed by the propagation, dominated by the domestic traveling patterns, to the less globally connected regions in the US. The model suggests that COVID-19 spread across the US in about 7 weeks, and that by the end of February many states were experiencing sustained SARS-CoV-2 local transmission. The median time for the onset of local transmission in coastal states is as early as February 2020, and it raises the question of what would the unfolding of the epidemic have looked like in the case of an earlier warning and issuing of travel restrictions to/from China. For this reason we have performed simulations of a counterfactual scenario in which the timeline of all the travel restrictions to/from mainland China and interventions is shifted one week earlier compared to the actual one. In Fig. 4E, we show the distribution of the probability that the US experiences sustained local transmission by a given date in both the counterfactual scenario and the real timeline of interventions. We define sustained national transmission as 100 new infections per day. Interventions implemented one week earlier amount to a proportional delay of the onset of local transmission in the continental US. In particular we find that in the real timeline intervention analysis a 50% cumulative probability of reaching the 100 locally generated transmissions mark is reached by February 2, 2020, compared to February 4, 2020, in the counterfactual scenario. This is in agreement with the evidence provided by several studies (*12, 47, 48*) that a considerable number of infections had already traveled from mainland China to international destinations before mid January, thus potentially seeding multiple epidemic outbreaks across the world, and leading to the international expansion of the COVID-19 epidemic, despite the mainland China travel ban. Our analysis however does not consider spontaneous behavioral changes that people might have adopted before the official national and local guidelines were announced. While certainly some individuals might have taken precautions as a result of the news from China, evidence from surveys of public concern from several countries in Europe suggest that in late February only a very limited fraction of people considered COVID-19 as a concrete threat (*49*). Our model also does not contain any calibration or constraint on the trajectory of the outbreak in the months of March and April. We provide this analysis in the SM showing the consistency of the results.

## Discussion

Our study characterizes the cryptic transmission phase during which SARS-CoV-2 spread largely undetected in the US. The results suggest that the first sustained local transmission chains took place as early as mid January, and by the end of February the infection was spreading to many other domestic locations. This timeline is shifted several weeks ahead with respect to the detection of cases in surveillance data. This is consistent with the fact that in January and February no country had the capacity to do mass testing. Countries adopted a policy of testing symptomatic individuals with a travel history linked to China, thus, generally missing the cryptic transmission occurring domestically. We find that the order in which the virus initially progressed across states according to our model is highly correlated with the official record. The model highlights that the geographical heterogenities in the observed spreading patterns are explained by the features of the air transportation network and population distributions. The results also indicate that the source of introduction of SARS-CoV-2 infections into the US changed substantially and rapidly through time. While early importations were from international sources, most introductions occurred during February and March 2020. Our results indicate that many states were seeded from domestic sources rather than international. The presented results could be of potential interest in combination with sequencing data of SARS-CoV-2 genomes to reconstruct in greater detail the early epidemic history of the US COVID-19 epidemic. The estimated SARS-CoV-2 importation pattern and the cryptic transmission phase dynamic are of potential use when planning and modelling public health policies in the context of international travel.

## Data Availability

Proprietary airline data are commercially available from Official Aviation Guide (OAG) and IATA databases.  The GLEAM model is publicly available at http://www.gleamviz.org/.

## Acknowledgements

M.E.H. acknowledges the support of the MIDAS-U54GM111274. S.M. and M.A. acknowledge support from the EU H2020 MOOD project. C.G. and L.R. acknowledge support from the EU H2020 Icarus project. M.C. and A.V. acknowledge support from Google Cloud and Google Cloud Research Credits program to fund this project. The findings and conclusions in this study are those of the authors and do not necessarily represent the official position of the funding agencies, the National Institutes of Health, or the U.S. Department of Health and Human Services.

## Author Contributions

Author contributions: J.T.D., M.C., N.P. and A.V. designed research; M.C., J.T.D., N.P., K.M., A.P.P, M.A., N.E.D., C.G., M.L., S.M.,L.R., K.S., X.X., M.E.H., I.M.L., C.V., and A.V. performed research; M.C., J.T.D., N.P., A.P.P., K.M. and A.V. analyzed data; and M.C., J.T.D., N.P., K.M., A.P.P., M.A., N.E.D., C.G., M.L., S.M., L.R., K.S., X.X., M.E.H., I.M.L., C.V., and A.V. wrote and edited the paper.

